# Exploratory and confirmatory factor analysis of the Cognitive Social Capital Scale in a Colombian sample

**DOI:** 10.1101/2020.09.22.20199869

**Authors:** Adalberto Campo-Arias, Carmen Cecilia Caballero-Domínguez, John Carlos Pedrozo-Pupo

## Abstract

During the last two decades, the concept of social capital has been used with increasing frequency in health sciences due to the direct and indirect relationships between social capital and populations’ physical and mental health. Therefore, it is necessary to build an instrument to quantify this concept confidently and reliably. The study aimed to perform exploratory and confirmatory factor analyses on a seven-item scale to measure social capital in adults of Colombia’s general population. An online validation study was done, including a sample of 700 adults aged between 18 and 76 years; 68% were females. Participants completed a seven-item scale called the Cognitive Social Capital Scale (CSCS). Cronbach’s alpha and McDonald’s omega were computed to test internal consistency. Exploratory and confirmatory factor analyses were conducted to explore the dimensionality of the CSCS. The CSCS presented a low internal consistency (Cronbach’s alpha of 0.56 and McDonald’s omega of 0.59) and poor dimensionality. Then, a five-item version (CSCS-5) was tested. The CSCS-5 showed high internal consistency (Cronbach’s alpha of 0.79 and McDonald’s omega of 0.80) and a one-dimension structure with acceptable goodness-of-fit indicators. In conclusion, the CSCS-5 presents high internal consistency and a one-dimension structure to measure cognitive capital social in Colombian sample. It can be recommended for the measuring of social capital in the general Colombian population. Further research should corroborate these findings on pencil and paper applications and explore other reliability and validity indicators.

## INTRODUCTION

In the last two decades, there has been an increasing application of the concept of social capital in various fields of public health; therefore, social capital is frequently used in the health sciences^1^. However, social capital is a controversial concept with extensive debates on the definition and objective measurement^2^. The most critical and practical limitation is the measurement of social capital in different clinical and epidemiological studies^3^.

The construction of an instrument to quantify social capital is a complex process due to the nature of the concept^4^. Nevertheless, it is necessary to have a reliable measurement of the construct in clinical and epidemiological studies^5^. Social capital is widely understood as the structural, relational, and cognitive characteristics of social interactions that facilitate concerted or coordinated actions and collective learning^4^. It can also define how the institutions, relationships, attitudes, and values mediate interactions among citizens and support economic and social development^6^. Social capital is understood to refer to the resources to which citizens and groups have access through constructed social networks^1^. Finally, it is currently defined as the set of direct or indirect resources results of social interaction between people and groups^7^. Then, some authors recognize three types of social capital structural, relational, and cognitive. While others only recognize two types of social capital, they make synonyms cognitive and relational social capital for the common elements between both concepts^8^.

According to the context, social capital can be divided into three types, bonding, bridging, and linking^2,9,10^. Bonding social capital refers to the resources available within the network for social relationships between individuals with similar characteristics, such as age, social class, or ethnicity/race^10^. Bridging social capital describes the social resources that people with different sociodemographic characteristics can access^1^. Linking social capital refers to networks of trust and respect that connect people and groups through formal institutions with authority or power^9^.

Also, there is one pair of types of social capital, which is relevant to health research. First, cognitive social capital refers to the perception of trust, reciprocity, honesty, truthfulness, and support from other people^8,11,12^. The second, structural social capital, refers to formal structures in which citizens can develop ties and social networks and participate in civic associations and events^1^. These concepts are particularly relevant due to the direct and indirect relationships between cognitive and structural social capital and the physical and mental health of different human groups^13,14^. For instance, De Silva et al^13^ reported an inverse association between cognitive social capital and common mental disorders, anxiety, and depression; likewise, the authors observed a moderate inverse relationship between cognitive social capital and mental disorders in children. Similarly, Riumallo-Herl et al^15^ reported that in people with diabetes or high blood pressure, the perception of physical health was inversely related to the perception of social capital.

Social capital has been measured in different ways, not always validly and reliably, from the psychometric perspective. The questions or items of these instruments have only evaluated the authors’ validity of appearance. For that reason, the available measurements for social capital are inconclusive and unable to quantify accurately the complex dimensions raised in theory^2,12,13^. To date, several English and Chinese instruments have been designed; these measurement scales are made up of different numbers of items and purposes. Some psychometric tests of reliability and validity were carried out on these instruments: The General Capital Social Scale^16^, Bonding Social Capital Scale^17^, Personal Social Capital Scale^18^, Social Capital Investment Inventory^19^, and Trust Scale based on Social Capital in Spanish^20^. Also, Wang’s scale has a Spanish version^21^. Other sets of items have been used as scales without performing a formal process of exploring psychometric performance in other studies. Without intention, nomological validity was achieved by exploring correlations with other variables of interest. Martin et al^22^ concluded that elevated social capital, particularly in terms of reciprocity among neighbors, was related to the low risk of household food security. Sapag et al^23^ reported that neighborhood social cohesion, measured by trust and reciprocity, was related to better self-assessment of health within a low-income community in Santiago, Chile. Alvarado et al^24^ observed a negative correlation between scores for social capital and psychological distress anxiety and depression among Chilean workers. For further illustration, Holt-Lunstad et al^25^ reported that solid social relationships could reduce mortality risk by up to 50%.

In the present study, seven face validity items were chosen to measure cognitive social capital, which is most related to mental health^13,24^. Martin et al^22^ took these items as a proxy instrument for measuring the social capital of a scale developed by other researchers to quantify social cohesion and trust^26^. Martin et al^22^ did not report any psychometric indicators for the set of items. The current authors named the tool as “Cognitive Social Capital Scale” (CSCS). The CSCS has several questions in common with other instruments that explore cognitive social capital^1,2,11,12^. A validity test was done exploring internal consistency and using exploratory and confirmatory factor analysis. Internal consistency reliability of tests represents the average of the correlations between the scores of the scale’s items^27,28^. On the other hand, factor analysis is technical to assessment dimensionality, which is a mathematical way of testing if the items are distributed in the theoretically proposed factors or dimensions^29,30^.

It is necessary to have blunt instruments to measure social capital that shows an acceptable or excellent psychometric performance^12,31^. Validity and reliability of measurements are essential for health research’s internal validity^5^. With the repeated use of the same instrument, it is possible to make more precise comparisons of the findings in different studies and contexts^31^.

The present study aimed to assess the reliability (internal consistency) and validity (exploratory and confirmatory factor analyses) of the CSCS in adults of Colombia’s general population.

## MATERIALS AND METHODS

### Design

A validation analysis without external reference criteria was conducted. The study was nested in a cross-sectional study that included several measurement scales, including a version of the CSCS. Validation studies are also known as methodological studies, as they explore the usefulness of some measurements or quantification of concepts. There are criterion-referenced validation studies that are the best available objective measurement to test the performance of the scale. These objective measurements are rare in the measurement of social and psychological concepts. While studies without reference criteria use tests or statistical techniques to approximate the reliability and validity of the measurements, as in the current present study in which exploratory and confirmatory factor analyses and internal consistency were performed.

### Participants

Seven hundred people residing in Colombia completed the research questionnaire. This sample size is acceptable for carrying out exploratory and confirmatory factor analyses with a minimum acceptable error in the estimates made; in general, samples larger than 500 participants are recommended^32^. The participants’ ages were between 18 and 76 years (mean=37.1, SD=12.7). Other demographic characteristics, in frequencies and percentages, are presented in Table 1.

**Table 1.** Demographical characteristics of participants.

| Variable | n | % |
| --- | --- | --- |
| **Gender** |  |  |
| Female | 476 | 68.0 |
| Male | 224 | 32 |
| **Permanent couple** |  |  |
| Yes (Civil partnership or married) | 336 | 48.0 |
| No (Single, separated, or widowed) | 364 | 52.0 |
| **Income (Colombian status)** |  |  |
| Low (I, II or III) | 420 | 60.0 |
| High (IV, V, or VI) | 280 | 40.0 |
| **Education** |  |  |
| High school or less | 72 | 10.3 |
| College or higher | 628 | 89.7 |

### Instrument

The participants completed the CSCS. This scale explores the current perceptions about the relationship among neighbors. Each item gives four response alternatives from strongly disagree to agree strongly, rated from zero to three^22^. The translation and back-translation processes were carried out based on international recommendations to get a culturally adapted and linguistically equivalent translation^33,34^. Two independent bilingual persons translated the items from English to Spanish. There were few differences in these versions that were resolved by consensus. A third person translated the final Spanish version into English. There was a high agreement in words and linguistic equivalence. Caution was taken to avoid negative sentences that, in Spanish, tend to confuse the sense highly when answering^35^. Below are the items of the CSCS, they were headed with the phrase “In the block, residential complex, building, or neighborhood in which I live”:

1. “People around here are willing to help their neighbours.” (La gente está dispuesta a ayudar a los vecinos).
2. “This is a close-knit, or ‘‘tight’’ neighborhood where people generally know one another.” (Las personas son unidas y generalmente se conocen entre sí).
3. “If I had to borrow $30 in an emergency, I could borrow it from a neighbor.” (Si tuviera que pedir prestado $50.000, en caso de emergencia, podría pedírselo prestado a un vecino).
4. “People in this neighborhood generally do not get along with each other.” (La gente generalmente se lleva bien).
5. “People in this neighborhood can be trusted.” (Se puede confiar en las personas).
6. If I were sick I could count on my neighbors to shop for groceries for me.” (Si estuviera enfermo podría contar con mis vecinos para que hiciera algunas compras por mí).
7. “People in this neighborhood do not share the same values.” (Las personas comparten los mismos valores).

### Procedure

The information was collected by distributing an electronic questionnaire sent by email, WhatsApp, and Facebook to the researchers’ contacts. The period to respond was limited from March 30 to April 8, 2020. In online studies, the highest rate of response, around 30%, occurs in the first week and does not improve considerably with additional requests^36^. The questionnaire did not ask for the name or other identifying information of the responder. All the questions in the questionnaire were mandatory; consequently, the participants had to complete all the items. This obligation was implemented to avoid incomplete or lost data.

### Analysis of data

In the validation studies, the variables are the items that are part of the scale. Generally, the process of exploring item performance begins with the calculation of internal consistency. Internal consistency is a measure that summarizes the mean of the correlations between the items that are part of a measurement scale. It is a measure of reliability and, at the same, an indirect measure of validity. High internal consistency is a fundamental requirement of measurements; however, it does not guarantee good performance on all validity of tests^37^. For instruments under construction, internal consistency values of 0.60 may be acceptable, but, for more developed instruments, values between 0.90 and 0.95 are preferable^5,38^. Internal consistency was calculated with Cronbach^27^ alpha and McDonald^28^ omega coefficients. Cronbach’s alpha coefficient is the most informed of the reliability indicators; however, it has the main limitation that it starts from the assumption of tau equivalence. All items contribute in a similar way to the construct^37^. When the principle of tau equivalence is violated, McDonald’s omega is a more accurate estimator of reliability^28,37^. These coefficients can be misinterpreted when they are calculated for multidimensional instruments since they erroneously overestimate the internal consistency due to the number of items^37^. Currently, it is recommended to report at least two reliability indicators for one-dimensional measurements^38^.

Factor analysis is used to identify an underlying or latent factor in a set of items. The exploratory factor analysis is used to test the dimensionality of a scale, often misnamed construct validity, and attempt to mathematically demonstrate the theoretical dimensions of an instrument in the initial stages of construction of an instrument^39^. The confirmatory factor analysis is used to confirm a previously suggested structure with advanced statistical procedures^40^. A researcher can expect up to two dimensions for a seven-item instrument, ideally one^29,30,41,42^.

The factor analyses were performed to test the dimensionality of the CSCS using the maximum likelihood method. The Kaiser-Meyer-Olkin index (KMO)^43^, and Bartlett^44^ sphericity test were computed in the first step of the exploratory factor analysis. If the KMO is more significant than 0.60 and Bartlett’s chi-square shows a p-value less than 0.05, they indicate that the items do indeed group a latent factor and the exploration can continue, without guarantees of an utterly satisfactory finding. After, communality and factor loadings are observed, interpreted as other correlation coefficients, and indicate the magnitude of the relationship between the item and the factor^45^.

Besides, confirmatory factor analysis is used to confirm a previously suggested structure with advanced procedures of computing goodness-of-fit coefficients: Root Mean Square Error of Approximation (RMSEA) and the 90% confidence interval (90% CI), Comparative Fit Index (CFI), the index Tucker-Lewis (TLI) and Standardized Mean Square Residual (SMSR). In the best conditions, it is expected chi-square with a probability greater than 0.05, RMSEA and SMSR with values that are close to 0.06; and for CFI and TLI values greater than 0.89. At least three coefficients within the desirable values may be sufficient to accept that the analyzed data fit the theoretical model of the instrument evaluated^46^. The analysis was completed in the statistical program STATA 13.0^47^.

### Ethical issues

The research was endorsed by the research ethics board of a Colombian state university (Act 002 of the extraordinary session held on March 26, 2020). The research followed the ethical recommendations for research on human subjects following the Declaration of Helsinki^48^, and the Colombian legislation that disapproves the provision of any incentives to research participants^49^. The participants gave informed consent. Besides, the participants’ anonymity, respect for privacy, and handling of all the information recorded in the research questionnaire were guaranteed.

## RESULTS

### CSCS

The CSCS showed low indicators of internal consistency; the value of Cronbach’s alpha was 0.56 and McDonald’s omega of 0.69. The exploratory factor analysis showed that the seven items of CSCS could retain a latent factor, the coefficients were excellent, Bartlett’s chi-square of 1,224.3, df=21, p<0.01, and KMO index of 0.77. Nevertheless, item 1 (helping the neighbors) showed a negative loading value, and item 4 (getting along) presented a very low loading. Commonalities and loadings of the CSCS are presented in Table 2.

**Table 2.** Commonalities and loadings of the CSCS.

| Item | Commonality | Loading |
| --- | --- | --- |
| 1. To help their neighbors | 0.03 | -0.18 |
| 2. "Tight" neighborhood | 0.18 | 0.42 |
| 3. To borrow \$30 | 0.48 | 0.69 |
| 4. Get along with | 0.01 | 0.09 |
| 5. Trust | 0.53 | 0.73 |
| 6. If I were sick | 0.64 | 0.80 |
| 7. Share the same values | 0.47 | 0.69 |

Besides, confirmatory factor analysis showed that all goodness-of-fit indicators were suboptimum. Then, the hypothesis of the one-dimensional structure is rejected. Likewise, the possible performance of a two-dimensional structure was explored, and the results were unsatisfactory (these coefficients are omitted). The indicators for the one-dimension structure of the CSCS are presented in Table 3.

**Table 3.** Goodness-of-fit indicators for the CSCS and CSCS-5.

| Item | CSCS | CSCS-5 |
| --- | --- | --- |
| X<sup>2</sup> (df)* | 194.77 (14) | 35.14 (5) |
| X<sup>2</sup>/df | 13.91 | 7.03 |
| RMSEA (90% CI) [p] | 0.14 (0.12-0.15) [p<0.001] | 0.09 (0.07-0.12) [p=0.005] |
| CFI | 0.85 | 0.97 |
| TLI | 0.80 | 0.94 |
| SRMR | 0.08 | 0.03 |

\*p<0.01.

### CSCS-5

After deleting items 1 and 4, this five-item version (CSCS-5) presented high internal consistency values, Cronbach’s alpha was 0.79 and McDonald’s omega of 0.80. Table 4 presents more descriptive information on the CSCS-5, mean, standard deviation, corrected item correlation, and Cronbach’s total score and alpha if the item were omitted.

**Table 4.** Mean, standard deviation, correlation corrected with the total score, and Cronbach’s alpha with the omission of the item of the CSCS-5.

| Item | M | SD | CC<sup>1</sup> | Alfa de Cronbach<sup>2</sup> |
| --- | --- | --- | --- | --- |
| 2. "Tight" neighborhood | 1.39 | 0.88 | 0.37 | 0.81 |
| 3. To borrow \$30 | 1.32 | 1.01 | 0.59 | 0.75 |
| 5. Trust | 1.68 | 0.76 | 0.63 | 0.74 |
| 6. If I were sick | 1.54 | 0.86 | 0.68 | 0.71 |
| 7. Share the same values | 1.51 | 0.77 | 0.60 | 0.75 |

<sup>1</sup>Corrected correlation.
<sup>2</sup>If the item is deleted.

The exploratory factor analysis indicators suggested that the five retained items represented a latent factor or dimension, Bartlett’s chi-square showed 1,064.3, df=10, p<0.01, and KMO was 0.81. Confirmatory factor analysis for CSCS presented acceptable coefficient values for a one-dimension structure. Table 3 shows the goodness-of-fit indexes for the CSCS-5 and CSCS, and Table 5 summarises the commonalities and loadings of the CSCS-5.

**Table 5.**
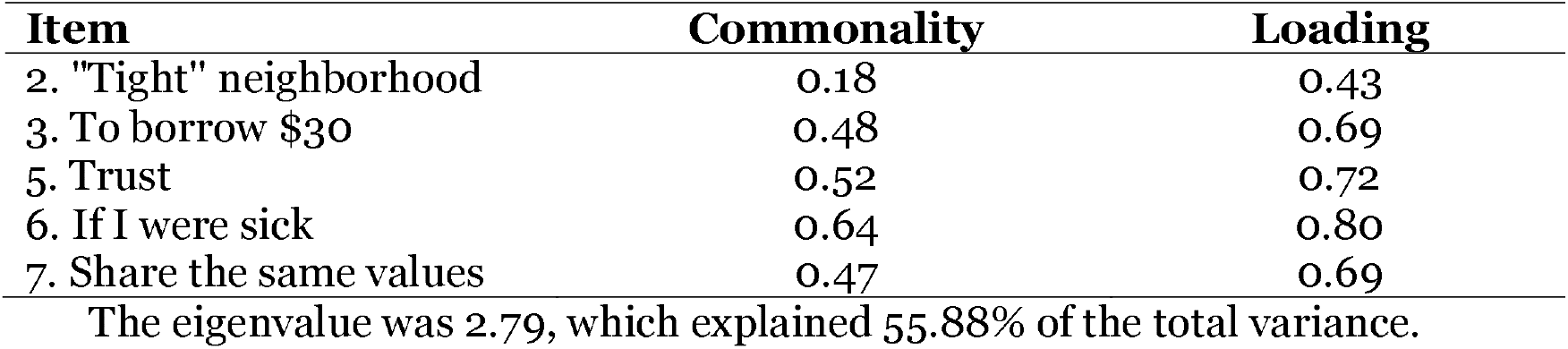
Commonalities and loadings of the CSCS-5.

| Item | Commonality | Loading |
| --- | --- | --- |
| 2. "Tight" neighborhood | 0.18 | 0.43 |
| 3. To borrow \$30 | 0.48 | 0.69 |
| 5. Trust | 0.52 | 0.72 |
| 6. If I were sick | 0.64 | 0.80 |
| 7. Share the same values | 0.47 | 0.69 |

## DISCUSSION

In this study, the psychometric performance of the CSCS and CSCS-5 is reported. Only the CSCS-5 shows good psychometric indicators of reliability and dimensionality. The CSCS-5 is a tool with adequate internal consistency and one-dimensional structure to assess cognitive social capital, that is, the perception of trust, reciprocity, and support that people have about other individuals and institutions^1,11,12^.

During the most recent decades, the interdisciplinary work between biomedical and social sciences makes necessary to guarantee the validity and reliability of the social variables’ measurements, usually approached qualitatively^31^. In general, one-dimensional instruments are expected to show three out of five favorable goodness-of-fit indicators^46^, and high internal consistency reliability^38^.

The CSCS-5 showed three out of five adequate goodness-of-fit coefficients for dimensionality. The most-reported coefficients are the Satorra-Bentler chi-square (with X^2^/df), the root of the mean square error of approximation (RMSEA), the Comparative Fit Index (CFI), the Tucker-Lewis Index (TLI), and the Standardized Mean Square Residual (SRMR). For the Satorra-Bentler chi-square is recommended a p-value higher than 0.05 (with X^2^/df<5); RMSEA, a value of around 0.06; CFI and TLI, values higher than 0.90; and SRMR, a value below 0.05^46^.

Likewise, the CSCS-5 achieved two excellent internal consistency indicators, Cronbach’s alpha of 0.79 and McDonald’s omega of 0.80. Measuring construct instruments should present high internal consistency reliability, with values between 0.70 and 0.95^38^. It is necessary to report an instrument’s internal consistency every time it is used to measure variables because it can vary significantly between samples from different populations^5^.

### Practical implications

The provision of the CSCS will facilitate new researches on the issue. The lack of short, reliable, and valid tools to assess cognitive social capital has been a problem for conducting considerable researches in the general population^2,12,13^. In health sciences, it is necessary to consider and evaluate cognitive social capital, which is most related to mental health outcomes; when the social determinants of health are increasingly taken into consideration, the physical and mental health of the population groups is the result of the complex interaction between individual, family, community, social, cultural and institutional factors^13^.

## Strengths and limitations

This study shows the performance of a short scale to quantify cognitive social capital using confirmatory factor analysis. This technic is a robust multivariate psychometric analysis that identifies latent factors underlying a set of items^41^. However, the technique does not overcome all the practical difficulties of such complex concepts as cognitive social capital^1,4^. The instruments must be adapted or modified as progress is made and the concepts to be measured are clarified^50^. Moreover, given the sample collection technique, the test-retest reliability of the scales could not be established^51^. This measurement is essential to guarantee the measurements’ total reliability with the instruments^5^. Similarly, the analysis of the device’s dimensionality and reliability in other languages is required to make valid comparisons of studies conducted in different languages^33,34^.

Future research should test the quality of this instrument by exploring other measures of validity: convergent with other instruments that quantify the same construct, divergent validity with an instrument that theoretically and empirically does not show any relationship with the construct of cognitive social capital, and different forms of nomological validity, that is, the relationship with other unrelated contexts; but, theoretically associated with cognitive social capital^5^. Besides, based on the item response theory, it is recommended to explore the differential functioning of the items to avoid that the instrument can make a biased measurement based on a characteristic utterly external to the instrument. Some traits of the participating population can be age, gender, or social or cultural background^52,53^. Unlike the classical theory, the item response theory is an alternative that allows identifying systematically biased response patterns from intuitive statistical analyses^54,55^.

In the same way, it is necessary to carry out other reliability measures, such as stability or test-retest^51^. Furthermore, sensitivity to change in repeated measurements over time^56^. It is necessary to observe the instrument’s performance in traditional pencil and paper measurements^57^. Finally, the validity and reliability of the CSCS-5 must be shown in different populations, according to age or other variables of interest^5^.

## Conclusions

It is concluded that the CSCS scale presents a one-dimensional structure that does not fit the data and low internal consistency. The CSCS-5 exhibits a better one-dimensional structure and internal consistency. It is necessary to corroborate the present findings in future online and face-to-face studies in other Colombian populations. It is vital to have a valid and reliable instrument to evaluate social capital in countries with significant inequalities, such as Colombia.

## Data Availability

The data will be shared with a justified request to the corresponding author.

## Notes

**Funding:** Universidad del Magdalena, Santa Marta, Colombia.

### Competing Interest Statement

The authors have declared no competing interest.

### Funding Statement

Universidad del Magdalena, Santa Marta, Colombia.

### Author Declarations

Research Ethics Board of the Universidad del Magdalena, Santa Marta, Colombia.

### Summary of Updates

This revised version expands the introduction with greater precision and conceptual clarity and gives more details of the process in the method.

